# Predicting intention to receive COVID-19 vaccine among the general population using the Health Belief Model and the Theory of Planned Behavior Model

**DOI:** 10.1101/2020.12.20.20248587

**Authors:** Liora Shmueli

**Affiliations:** Department of Management, Bar-Ilan University, Ramat-Gan, 52900, Israel.

**Keywords:** Vaccine acceptance, COVID-19, health belief model, theory of planned behavior

## Abstract

**Background:** A novel coronavirus (COVID-19) was declared a global pandemic by the World Health Organization (WHO) in March, 2020. Until such time as a vaccine becomes available, it is important to identify the determining factors that influence the intention of the general public to accept a future COVID-19 vaccine. Consequently, we aim to explore behavioral-related factors predicting intention to receive COVID-19 vaccine among the general population using the Health Belief Model (HBM) and the Theory of Planned Behavior (TPB) model.

**Methods:** An online survey was conducted among adults aged 18 years and older from May 24 to June 24, 2020. The survey included socio-demographic and health-related questions, questions related to the HBM and TPB dimensions, and intention to receive COVID-19 vaccine. Associations between questionnaire variables and COVID-19 vaccination intention were assessed by univariate and multivariate analyses.

**Results:** Eighty percent of 398 eligible respondents stated their willingness to receive COVID-19 vaccine. A unified model including HBM and TPB covariates as well as demographic and health-related factors, proved to be a powerful predictor of intention to receive COVID-19 vaccine, explaining 78% of the variance (adjusted R2 = 0.78). Men (OR=4.35, 95% CI 1.58–11.93), educated respondents (OR=3.54, 95% CI 1.44–8.67) and respondents who had received the seasonal influenza vaccine in the previous year (OR=3.31, 95% CI 1.22–9.00) stated higher intention to receive COVID-19 vaccine. Participants were more likely to be willing to get vaccinated if they reported higher levels of perceived benefits of COVID-19 vaccine (OR=4.49, 95% CI 2.79–7.22), of perceived severity of COVID-19 infection (OR=2.36, 95% CI 1.58–3.51) and of cues to action (OR=1.99, 95% CI 1.38–2.87), according to HBM, and if they reported higher levels of subjective norms (OR=3.04, 95% CI 2.15–4.30) and self-efficacy (OR=2.05, 95% CI 1.54–2.72) according to TPB. Although half of the respondents reported they had not received influenza vaccine last year, 40% of them intended to receive influenza vaccine in the coming winter and 66% of them intended to receive COVID-19 vaccine.

**Conclusions:** Providing data on the public perspective and predicting intention for COVID-19 vaccination using HBM and TPB is important for health policy makers and healthcare providers and can help better guide compliance as the COVID-19 vaccine becomes available to the public.

## Background

On December 31, 2019, a novel strain of coronavirus (COVID-19) was identified in Wuhan, China. In the following months, the outbreak spread all over the world and was declared a global pandemic by the World Health Organization (WHO) on March 11, 2020 [1]. COVID-19 is a highly contagious disease and according to the WHO, there have been more than 46 million confirmed cases of COVID-19, including more than a million deaths worldwide, a fifth of them in the United States alone, as of November, 2020 [2]. Along with the mortality, the pandemic has had an enormous economic impact, having caused the largest recession in history, including a high increase in unemployment levels. Economic assessments suggest that on average, each additional month of the crisis costs 2.5-3% of the global GDP [3].

At the time of writing this paper (October 2020), no vaccine to COVID-19 has become available yet. More than 150 countries engaged in generating a COVID-19 vaccine, which will probably be available only by the end of 2021 [4]. Until such time as a vaccine becomes available the WHO had strongly recommended a series of actions designed to flatten the infection curve. These include the use of medical masks, frequent hand washing with sanitizer or soap and maintaining social distancing [5]. Even when a vaccine becomes available, priority will be given to vulnerable groups in the population, according to their level of risk. Before a COVID-19 vaccine becomes available, it is important to understand the intentions, motivators and barriers that influence the intention of the general public to accept a future COVID-19 vaccine. Such understanding would help prepare intervention plans based on accessibility to the general public while targeting populations that show a tendency not to get vaccinated.

To date, only few studies have studied the intention of the public to get vaccinated once a COVID-19 vaccine becomes available. A study conducted in Europe, involving participants from Denmark, France, Germany, Italy, Portugal, the Netherlands, and the UK, demonstrated a high response rate of 74% [6]. A higher response rate of 86% was found in Australia [7], while a lower rate of 69% was found among adults in the United States [8]. In considering the factors associated with willingness to be vaccinated against COVID-19, one can divide them into demographic and health-related predictors and predictors based on theoretical behavior models.

### Demographic and health-related predictors

Recent studies addressing predictors of intention to receive COVID-19 vaccine have shown that significantly higher proportion of men were willing to get vaccinated (77.9%) than women (70.1%), especially among men above the age of 55 [6]. Likewise, individuals considering themselves at risk for the disease [9] and those who reported their healthcare provider would recommend they get vaccinated against COVID-19 [8] were more likely to self-report acceptability to receive COVID-19 vaccination. While only few studies have investigated willingness to receive a vaccine against COVID-19, many studies have investigated acceptance of influenza vaccine. In the present study, aimed at determining the willingness to receive a COVID-19 vaccine, we adopted some of the factors studied in the case of influenza vaccine.

The literature reports several dominant characteristics that describe patients who intended to get a flu vaccine. Males are apparently more willing to get vaccinated than are woman [10, 11] and older patients above the age of 65 are more willing to get vaccinated than are younger patients [12]. At the same time, more educated patients and those having high income levels are willing to get vaccinated [13, 14], as are those having chronic health conditions and who perceived their health to be less good [15].

Other characteristics, such as living alone with no partner or children and being unmarried, were negatively associated with the desire to get vaccinated [11].

### Predictors based on theoretical behavior models

Theoretical models of health beliefs and risk perception are essential tools for understanding the factors behind decision-making by assessing what motivates and inhibits people to adopt health-related behavior. The HBM is one of the most widely used models for examining the relationship between health behavior and the use of health services. This strategy seeks to explain and predict preventive health behavior in terms of certain belief patterns. The HBM has been widely used in the context of vaccination, particularly influenza vaccination [10, 16, 17]. According to the HBM model, the intention to get an influenza vaccine depends on a number of factors, including perceived susceptibility and perceived severity, which reflects perception of the threat, as well as perceived benefits and perceived barriers as a function of outcome expectation and cues to action. Perceived susceptibility refers to individual perception regarding the chance of being infected by influenza. At one end of the scale one finds individuals who deny the possibility of infection, while at the other end one finds people who feel the danger of infection. In previous studies, this predictor was found to be a significant predictor for refusing a vaccine (43.2%), of patient perception of not being at risk for influenza, bias with a sense of disease resistance, and a low chance of the individual getting sick.

Perceived severity refers to the individual’s belief as to difficulties that the disease may create medically and socially, for example, pain, missing workdays, etc. Perceived barriers refer to the individual’s perceived negative aspects related to the action of getting vaccinated, such as expenses, physical pain, psychological considerations or a logistic lack of access [11, 18]. Cues to action is the last predictor that completes the behavioral change proposed in the original HBM, and include the presence of internal or external incentives that serve to motivate vaccination, such as information from the mass media or a doctor who recommends taking the vaccine [19].

TPB is another theoretical model used to predict an individual’s behavior in terms of intention to get vaccinated. According to the TPB model, the intention to get an influenza vaccination depends on a number of predictors, including the attitude towards the vaccine, subjective norms for carrying out vaccination, and perception of behavioral control (PBC) of vaccination. Self-efficacy for vaccination is another predictor that was added to the original model, as it has been proven that a distinction must be made between perception of control of behavior and self-efficacy. Self-efficacy was found to be the most important predictor for health behavioral intention [20, 21]. A few recent studies have combined the TPB and HBM approaches to identify health-related behaviors and intention to receive influenza vaccine among the general public [18, 22, 23].

In the context of COVID-19, several health beliefs have also been correlated with vaccine acceptability. Study participants who reported higher levels of perceived likelihood of getting a COVID-19 infection in the future and who perceived the severity of COVID-19 infection were more likely to be willing to get vaccinated [8]. The perceived benefit construct in the HBM was also found to be significant in predicting acceptance of the vaccine [24]. We are not aware of any study conducted on predicting COVID-19 vaccine acceptability based on the TPB model, although studies have used this model in the context of linking COVID-19 with preventive behaviors (e.g. social distancing, washing hands, etc.) [25, 26]. We are also not aware of any study in which both models were used to identify the factors of the general public’s willingness to receive a COVID-19 vaccine.

The aims of the present study were to investigate attitudes and beliefs of the general public regarding a future COVID-19 vaccination, and to identify the determining factors, motivators and barriers leading to the decision to receive vaccination or not, including factors adopted from the case of influenza, together with the combined use of the HBM and TPB models.

## Methods

### Study participants and survey design

#### Design

We conducted a cross-sectional national anonymous web-based survey using an electronic questionnaire, distributed via online social platforms (Google, Facebook and WhatsApp) among the general population. The survey was conducted between May 24 and June 26, after the Israeli government announced a variety of restrictions in May, 2020. These strict restrictions, including lockdown, obligation to wear a mask, etc., were decreed during March-April, following the proclamation of COVID-19 as a global pandemic. At that time, the COVID-19 vaccination was in development, according to the WHO, with 26 different vaccines in the human trial phase [4].

#### Process

Before distributing the questionnaire, experts validated the content and the questionnaire was pilot-tested. At the beginning of the questionnaire form, the respondents were informed that their participation was voluntary, and they confirmed consent to participate in the research. The interviewers followed a pre-defined closed-end protocol. We conducted a sample of the general Israeli adult population. The inclusion criterion was being 18 years of age and older and consent to participate.

#### Ethical considerations

The study was approved by the Ethics Committee for Non-clinical Studies at Bar Ilan-University in Israel.

#### Questionnaire

The following sections describe the dependent and independent variables and their operationalization in this study. Health belief measures were adopted from another study based on the HBM and TPB models [27, 28].

The parameters comprising the study measurements were used to build the conceptual model (see Figure 1) and are described below and in Table 1.

**Table 1.** Items, response scales and internal consistency for assessing measures of 2 models: HBM, TPB.

| Model | Measures | Items | Mean | Std | a |
| --- | --- | --- | --- | --- | --- |
| **HBM** | Perceived susceptibility | I believe that if I do not get vaccinated, the likelihood of me getting infected with corona will increase | 4.58 | 1.59 | 0.83 |
|  |  | I believe that if I do not get vaccinated, the likelihood of my family and relatives getting infected in Corona will increase | 4.50 | 1.55 |  |
|  | *Perceived severity | Even if I will get infected with COVID-19 I do not think it will cause me significant suffering or complications | 3.74 | 1.66 | 0.73 |
|  |  | Even if I get infected with COVID-19, the likelihood of recovering from the disease is very high | 2.63 | 1.38 |  |
|  | Perceived Benefits | I believe that COVID-19 vaccine will have high efficacy in preventing significant suffering and complications of the disease | 4.87 | 1.40 | 0.87 |
|  |  | I believe that if I get vaccinated against COVID-19 the risk of getting infected with the disease or infecting others will decrease | 4.97 | 1.39 |  |
|  | Perceived barriers | Getting vaccinated is expensive, requires time and effort | 2.44 | 1.53 | - |
|  | Cues to action | The chances of me getting vaccinated against COVID-19 will increase if opinion leaders on social media express support for the benefit of the vaccine | 2.98 | 1.94 | 0.79 |
|  |  | The chances of me getting vaccinated against COVID-19 will increase if friends and family express support for the benefit of the vaccine | 3.63 | 1.83 |  |
|  |  | The chances of me getting vaccinated against COVID-19 will increase if official guidelines from the Ministry of Health are published | 4.44 | 1.72 |  |
|  |  | The chances of me getting vaccinated against COVID-19 will increase if my GP recommends me | 4.29 | 1.77 |  |
|  |  | If my workplace takes care of vaccinating the workers against COVID-19, I will vaccinate | 4.48 | 1.90 |  |
|  | Health motivation | I exercise as recommended for my age | 3.90 | 1.70 | 0.75 |
|  |  | I make sure to eat a healthy and varied diet | 4.24 | 1.39 |  |
| **TPB** | Attitude | Getting vaccinated is a tedious process that requires time and effort | 2.44 | 1.53 | - |
|  | Subjective norms | Most of my friends will support the COVID-19 vaccine | 4.68 | 1.28 | 0.86 |
|  |  | If I tell my friends and relatives that I intend to get vaccinated against COVID-19 when a vaccine is available, they will respond positively | 4.99 | 1.23 |  |
|  | PBC | If I am offered a vaccine against COVID-19 for the coming winter, I'm sure I'll be vaccinated, and this decision is entirely up to me. | 5.05 | 1.29 | - |
|  | * Self-efficacy | If I take all the necessary precautions (disinfection of hands, etc.) I do not need to be vaccinated against corona | 4.87 | 1.33 | - |

a Cronbach indicates the internal consistency: **HBM** a= 0.77 **TPB** a= 0.60
Items Response scale: 1-6 agreement
\* Negative items were reverse scored.

**Figure 1.**
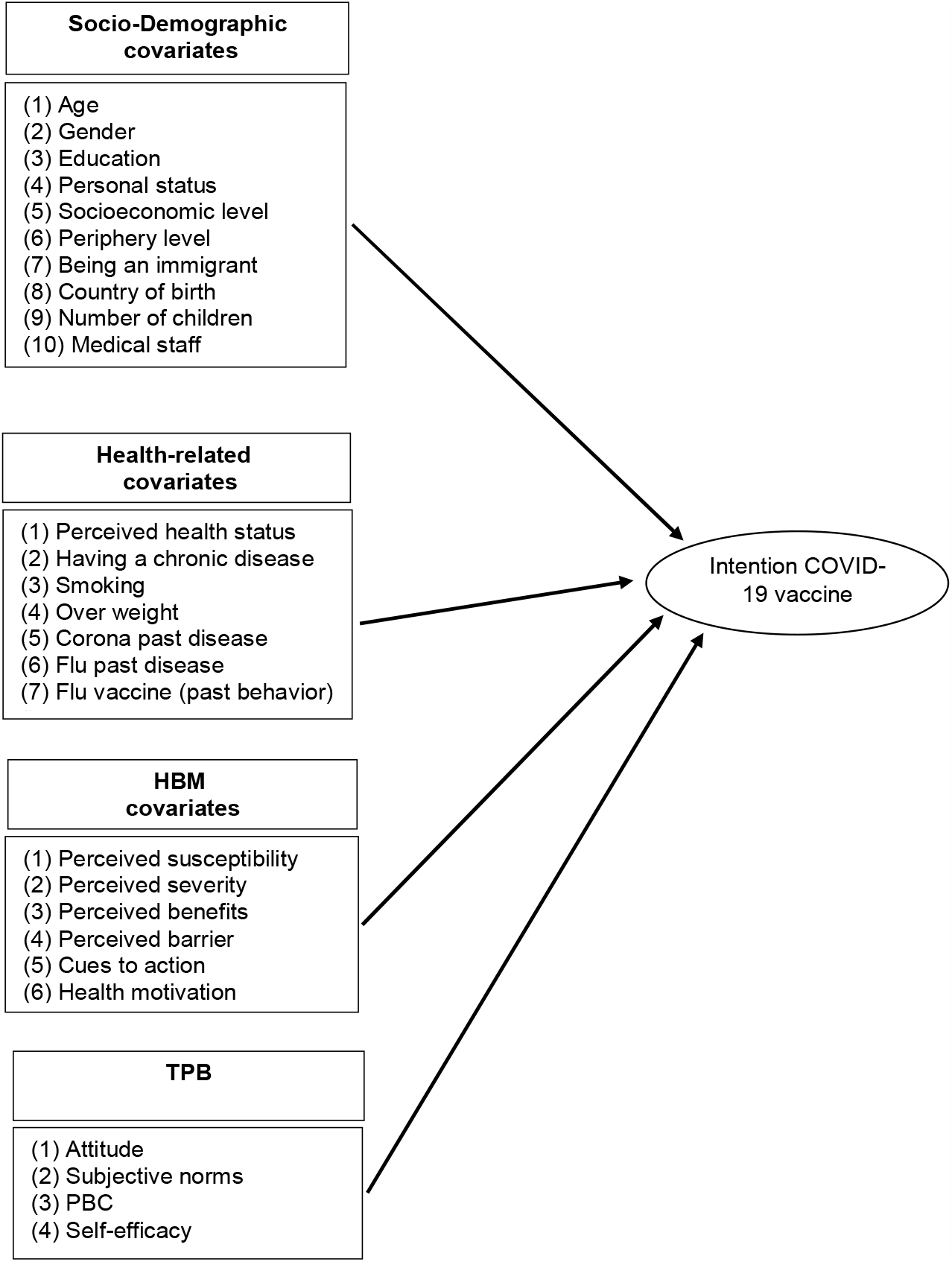
Conceptual Framework for the hypothesized predictors of intention to receive COVID-19 vaccine

The questionnaire consisted of the following sections: (1) socio-demographic covariates; (2) health-related covariates (3); HBM covariates (4) TPB covariates; (5) intention to receive a future COVID-19 vaccine; and (6) intention to receive an influenza vaccine. Overall, the questionnaire consisted of 45 questions and took less than 10 minutes to complete.

#### Variables and measurements

The dependent variable was the intention to a receive a future COVID-19 vaccine, measured by a one-item question on a 1-6 scale (1 - not appropriate at all, 6 - very appropriate).

1. The socio demographic covariates were: (1) age; (2) gender; (3) education level; (4) personal status (in partnership or not; with or without children); (5) socio-economic level, based on the Israeli Central Bureau of Statistics scale; (6) periphery level, defined by residential area; (7) being an immigrant (defined as immigration to Israel after 1989); (8) number of children; and (9) being medical staff or not.
2. The health-related covariates were: (1) perceived health status; (2) having a chronic disease; (3) smoking; (4) being over-weight; (5) past episodes of COVID-19; (6) past episodes of influenza; and (7) having received influenza vaccine last year (i.e., past behavior).
3. The HBM covariates were: (1) perceived susceptibility (included two items, Cronbach α =0.83); (2) perceived severity (included two items, Cronbach α =0.73); (3)perceived benefits (included two items, Cronbach α =0.87); (4) perceived barriers (included one item); (5) cues to action (included five items, Cronbach α =0.79); (6) health motivation (included two items, Cronbach α =0.75), a dimension added to the original model. Not many studies include this consideration as a model variable [21].
4. The TPB covariates were: (1) attitude (included one item); (2) subjective norms (included two items, Cronbach α=0.86); (3) PBC (Perceived Behavioral Control) and (4) self-efficacy. The last two covariates included one item each, and correspond to dimensions added to the original model [21].

Items in the HBM and TPB models were measured on a 1-6 scale (1-not appropriate at all, 6 - very appropriate). Negative items were reverse-scored. Scores for each item were averaged to obtain each of the HBM- and TPB-independent categories.

#### Reliability of the questionnaire

A Cronbach α internal reliability method revealed the internal consistency of HBM was Cronbach α=0.77 and of TPB model was Cronbach α =0.60 (Table 1).

## Statistical analyses

The data from the electronic questionnaires were imported into the SPSS 26 software and were identified by code alone. Data processing and analysis was done using SPSS 26 software. To test the reliability of HBM and TPB measures, Cronbach’s α test was used. To describe the study population characteristics, the following methods of descriptive statistics were used: frequencies, percentages, averages and standard deviations.

Relationships between dependent and independent variables were examined by univariate analyses, using either t-tests on independent samples, one-way Anova tests or x2 tests, depending on the characteristics of the examined variable.

To investigate determinants of intention to receive a COVID-19 vaccine, a hierarchical multiple logistic regression was performed. The intention to receive a COVID-19 vaccine was used as the dependent variable, where the original 6 categories variable (a scale from 1-6; 1 - very appropriate, 6 - not appropriate at all), was transformed to a binary variable (1 - intends to get vaccinated, 0 - does not intend to get vaccinated). With regard to the independent variables, only variables that were found in the univariate analyses to be significantly correlated (p < .05) with intention to receive the COVID-19 vaccine were included in the regression. These variables were divided into four blocks. Socio-demographic variables were entered into the first block, health-related factors were entered into the second block, followed by key variables from the HBM and the TPB models which entered into the third and fourth blocks, respectively.

## Results

### Participants Characteristics

Descriptive characteristics of the respondents are provided in Table 2. Overall, 398 respondents completed the survey, 60% of whom were female (n=238). The average age of those included in the sample was 42.9 years (SD= 14.7), with half of the participants aged 18-39 years. The majority of those included hold an academic degree (n=295) and most live with a partner (77%). 16.6% of respondents (n=66) stated that they suffer from at least one chronic disease, most suffering from hypertension (41.5%) or Diabetes mellitus (20.7%). A third of the respondents were overweight (n=128). Only 2% of participants indicated previous COVID-19 infection.

**Table 2:** Characteristics of respondents by intention to get covid-19 vaccine (n=398)

|  | All subjects<br>(n=398) |  | Not Intent to<br>vaccine covid-19<br>N=78 (20%) |  | Intent to<br>vaccine covid<br>N=320 (80%) |  | p-value |
| --- | --- | --- | --- | --- | --- | --- | --- |
| Sociodemographic | N | (%) | N | (%) | N | (%) |  |
| Age group |  |  |  |  |  |  |  |
| 18 thru 39 | 202 | (50.8%) | 48 | (23.8%) | 154 | (76.2%) | 0.031* |
| 40 thru 64 | 153 | (38.4%) | 27 | (17.6%) | 126 | (82.4%) |  |
| 65+ | 43 | (10.8%) | 3 | (7.0%) | 40 | (93.0%) |  |
| Gender |  |  |  |  |  |  |  |
| Male | 160 | (40.2%) | 18 | (11.3%) | 142 | (88.8%) | 0.001* |
| Female | 238 | (59.8%) | 60 | (25.2%) | 178 | (74.8%) |  |
| Educational level |  |  |  |  |  |  |  |
| Non-academic | 103 | (25.9%) | 37 | (35.9%) | 66 | (64.1%) | <0.001* |
| Academic | 295 | (74.1%) | 41 | (13.9%) | 254 | (86.1%) |  |
| Personal status-<br>partnership |  |  |  |  |  |  |  |
| Living with a partner | 306 | (76.9%) | 62 | (20.3%) | 244 | (79.7%) | 0.543 |
| Not living with a partner | 92 | (23.1%) | 16 | (17.4%) | 76 | (82.6%) |  |
| Personal status-living<br>with a child |  |  |  |  |  |  |  |
| Living with a chilled | 255 | (64.1%) | 52 | (20.40%) | 203 | (79.60%) | 0.549 |
| Not living with a chilled | 143 | (35.9%) | 26 | (18.20%) | 117 | (81.80%) |  |
| Socioeconomic level |  |  |  |  |  |  |  |
| Low | 31 | (7.8%) | 6 | (19.4%) | 25 | (80.6%) | 0.482 |
| Medium | 170 | (42.7%) | 37 | (21.8%) | 133 | (78.2%) |  |
| High | 191 | (48.0%) | 32 | (16.8%) | 159 | (83.2%) |  |
| Peripheral level |  |  |  |  |  |  |  |
| Periphery | 33 | (8.3%) | 7 | (21.2%) | 26 | (78.8%) | 0.779 |
| Intermediate | 170 | (42.7%) | 35 | (20.6%) | 135 | (79.4%) |  |
| Central | 190 | (47.7%) | 34 | (17.9%) | 156 | (82.1%) |  |
| Immigration |  |  |  |  |  |  |  |
| New Immigrant>89 | 28 | (7.0%) | 5 | (17.9%) | 23 | (82.1%) | 0.8 |
| Native-born and<br>established immigrants | 370 | (93.0%) | 73 | (19.7%) | 297 | (80.3%) |  |
| Number of Children |  |  |  |  |  |  |  |
| No childs | 97 | (24.4%) | 19 | (19.6%) | 78 | (80.4%) | 0.638 |
| 1-2 childs | 152 | (38.2%) | 33 | (21.7%) | 119 | (78.3%) |  |
| 3 childs | 112 | (28.1%) | 18 | (16.1%) | 94 | (83.9%) |  |
| 4+ childs | 36 | (9.0%) | 8 | (22.2%) | 28 | (77.8%) |  |
| Medical staff |  |  |  |  |  |  |  |
| yes | 44 | (11.1%) | 7 | (15.9%) | 37 | (84.1%) | 0.5 |
| no | 354 | (88.9%) | 71 | (20.1%) | 283 | (79.9%) |  |
| Health related characteristics |  |  |  |  |  |  |  |
| Chronic Disease |  |  |  |  |  |  |  |
| No chronic disease | 332 | (83.4%) | 70 | (21.1%) | 262 | (78.9%) | 0.023* |
| Chronic disease | 66 | (16.6%) | 8 | (12.1%) | 58 | (87.9%) |  |
| **Smoking** |  |  |  |  |  |  |  |
| Yes | 44 | (11.1%) | 13 | (29.5%) | 31 | (70.5%) | 0.078 |
| No/quitted | 349 | (87.7%) | 64 | (18.3%) | 285 | (81.7%) |  |
| **Over weight** |  |  |  |  |  |  |  |
| Yes | 128 | (32.2%) | 14 | (10.9%) | 114 | (89.1%) | <0.001* |
| No | 247 | (62.1%) | 62 | (25.1%) | 185 | (74.9%) |  |
| **Corona past disease** |  |  |  |  |  |  |  |
| Yes | 8 | (2.0%) | 2 | (25.0%) | 6 | (75.0%) | 0.693 |
| No | 371 | (93.2%) | 72 | (19.4%) | 299 | (80.6%) |  |
| **Flu disease last year** |  |  |  |  |  |  |  |
| yes | 43 | (10.8%) | 11 | (25.6%) | 32 | (74.4%) | 0.295 |
| no | 355 | (89.2%) | 67 | (18.9%) | 288 | (81.1%) |  |
| **Flu vaccine last year** |  |  |  |  |  |  |  |
| No | 205 | (51.5%) | 69 | (33.7%) | 136 | (66.3%) | <0.001* |
| yes | 193 | (48.5%) | 9 | (4.7%) | 184 | (95.3%) |  |
| **Perceived health status** |  |  |  |  |  |  |  |
| Very good | 295 | (74.1%) | 64 | (21.7%) | 231 | (78.3%) | 0.079 |
| Good | 89 | (22.4%) | 14 | (15.7%) | 75 | (84.3%) |  |
| Not so good | 14 | (3.5%) | 0 | (0.0%) | 14 | (100.0%) |  |

Note: Percentages of **Not Intent** to vaccine covid vs. **Intent** to vaccine covid are calculated as valid % per each row (i.e., each row sums up to 100%, without missing values).
\* $p < 0.05$

### Willingness to receive COVID-19 and influenza vaccines

Overall, 80% of participants were willing to receive COVID-19 vaccine (n=320). 48% of participants (n=193) reported having received influenza vaccine in the previous season.

Here, the rates reported for individuals aged 65 and above were significantly higher than for younger respondents aged 18-39 years (77% vs. 43%, p < 0.05). Although 52% (n=205) reported having decided not to receive influenza vaccine in the previous season, 40% of them indicated that they would be willing to get influenza vaccine in the coming winter and 66% of them reported they intended to get COVID-19 vaccine, once available.

### Univariate analyses

Results of the univariate analyses between socio-demographic and health-related variables and willingness to get vaccinated against COVID-19 are reported in Table 2.

The covariates that were found to have a statistically significant effect (p<0.05) on intention to receive COVID-19 vaccine were age group, gender, educational level, suffering from a chronic disease, being over-weight and having received influenza vaccine in the previous season. Covariates that were not found to be statistically significant include personal status, immigration, periphery level, socio-economic level, number of children, being medical staff, smoking, past episodes of COVID-19 or influenza, or perceived health status.

Results of the univariate analyses between HBM and TPB variables and willingness to get vaccinated against COVID-19 are reported in Table 3. Specifically, Table 3 shows the mean values of HBM and TPB covariates as values on a 1-6 agreement scale, reflecting the intention to receive COVID-19 vaccine. The results in Table 3 indicate that according to HBM, those who intend to get COVID-19 vaccine, on average, perceived COVID-19 to be a more serious illness than those who did not intend to take the vaccine. The former group was more susceptible to illness, perceived a higher risk of infection, perceived more benefits from vaccination, and had higher levels of cues to action. There was no significant difference between the two groups in terms of perceived barriers and health motivation. According to the TPB model, those who intend to get COVID-19 vaccine, on average, reported higher levels of subjective norms than those who did not intend to take the vaccine. The former group also reported higher levels of self-efficacy regarding the vaccine. There was no significant difference between the two groups in terms of attitude and PBC.

**Table 3.** Univariate analyses between HBM and TPB variables and willingness to get vaccinated against COVID-19

|  | DO not-intend to get vaccinated<br>(n= 78) |  | Intend to get vaccinated<br>(n= 320) |  | t-test | P value<br>(two-tail) |
| --- | --- | --- | --- | --- | --- | --- |
| **HBM covariates** | Mean (SD) |  | Mean (SD) |  |  |  |
| Perceived Susceptibility | 2.89 | (1.54) | 4.94 | (1.12) | -11.09 | 0.00 |
| Perceived Severity | 2.48 | (1.24) | 3.36 | (1.33) | -5.28 | 0.00 |
| Perceived Benefits | 3.10 | (1.44) | 5.37 | (0.79) | -13.41 | 0.00 |
| Perceived Barriers | 2.55 | (1.59) | 2.42 | (1.51) | 0.70 | 0.48 |
| Cues to action | 2.51 | (1.33) | 4.16 | (1.26) | -10.25 | 0.00 |
| Health motivation | 4.03 | (1.51) | 4.08 | (1.36) | -0.32 | 0.75 |
| **TPB covariates** | 2.55 | (1.59) | 2.42 | (1.52) |  |  |
| Attitude | 3.49 | (1.36) | 5.16 | (0.85) | 0.70 | 0.48 |
| Subjective norms | 4.91 | (1.51) | 5.08 | (1.24) | -10.42 | 0.00 |
| PBC | 3.53 | (1.44) | 5.20 | (1.06) | -0.95 | 0.35 |
| Self-efficacy | 2.89 | (1.54) | 4.94 | (1.12) | -9.67 | 0.00 |

COVID-19 vaccination intention measured by the item: “I want to get vaccinated against the COVID-19 virus soon in case there is a vaccine available”, on a 1-6 agreement scale
HBM and TPB Items Response scale: 1-6 agreement

### Factors associated with intention to receive COVID-19 vaccine

Our first model, which included HBM variables as well as demographic and health-related factors (Table 4; model 1), explained 74% of the variance in intention to receive COVID-19 vaccine (adjusted R2 = 0.74). The most important components of the hierarchical regression were the HBM dimensions, which added 45% to the explained variance, on top of the 29% explained by the demographic and health-related characteristics. According to this model, two demographic variables, gender and education, were associated with intention to get vaccinated against COVID-19. Men intended to receive COVID-19 vaccine more than woman (OR=4.35, 95% CI 1.58– 11.93) and educated respondents intended to receive COVID-19 vaccine more than non-educated respondents (OR=3.54, 95% CI 1.44–8.67). Only one health-related variable, i.e., having received influenza vaccine last year, was a significant predictor. Respondents who had received the seasonal influenza vaccine in the previous year were 3.3-fold significantly more likely to intend to get vaccinated against COVID-19, as compared with those who had not received the seasonal influenza vaccine in the previous year (OR=3.31, 95% CI 1.22–9.00).

**Table 4:**
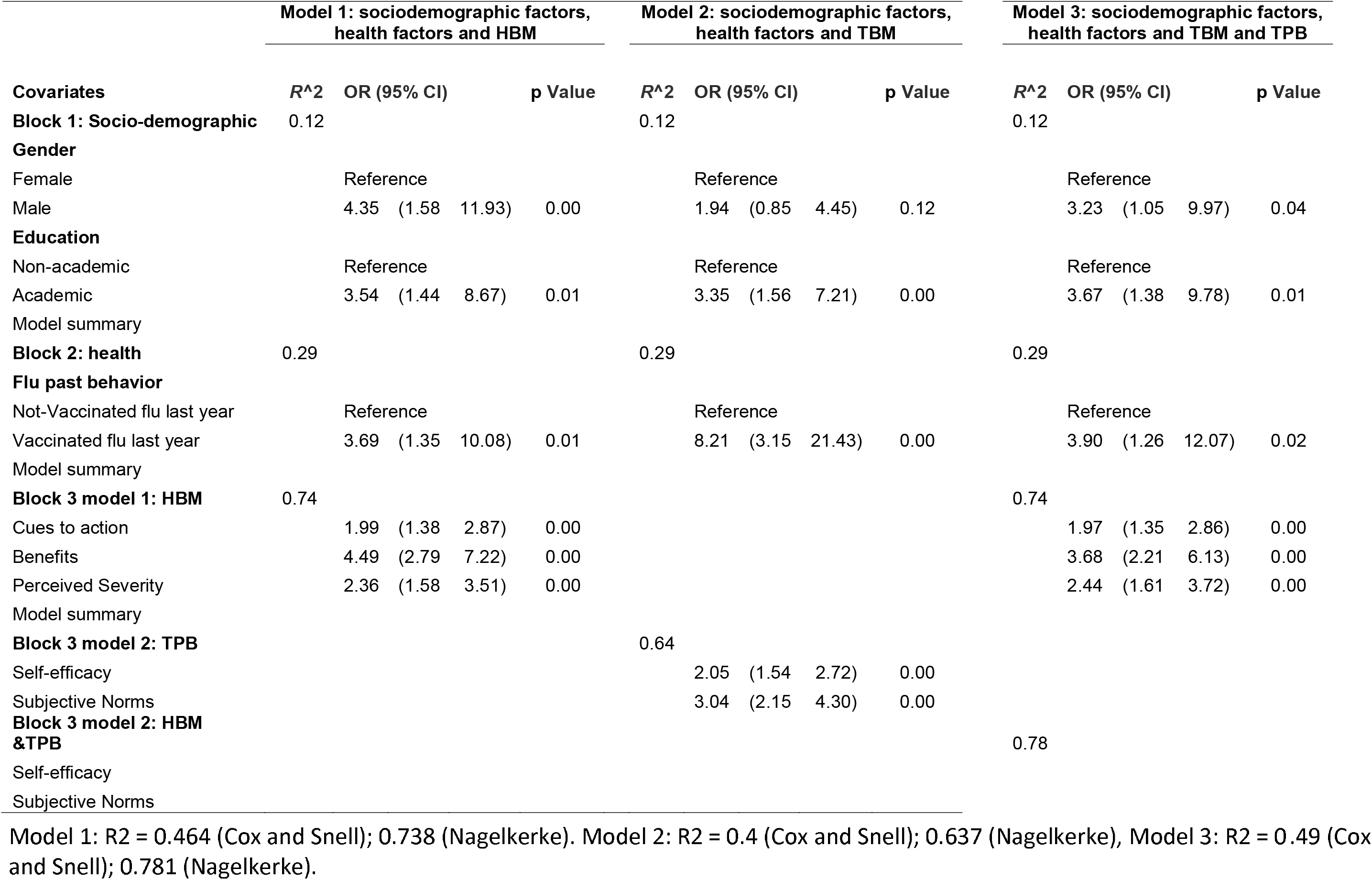
Hierarchical multiple logistic regression analysis-predictors of a covid-19 vaccine intention (n=398)

From the HBM, perceived benefits, cues to action, and perceived severity were significant predictors of intention to get vaccinated against COVID-19. Two perceived benefits, namely, “COVID-19 vaccine will have high efficacy in preventing significant suffering and complications of the disease” and “I believe that if I get vaccinated against COVID-19, the risk of getting infected with the disease or infecting others will decrease”, were significant predictors of intention to receive COVID-19 vaccine (OR=4.49, 95% CI 2.79–7.22); Two perceived severity statements, namely, “Even if I will get infected with COVID-19, I do not think it will cause me significant suffering or complications” and “Even if I get infected with COVID-19, the likelihood of recovering from the disease is very high” were also significant predictors of intention to get vaccinated against COVID-19 (OR=2.36, 95% CI 1.58–3.51). Finally, five cues to action: “The chances of me getting vaccinated against COVID-19 will increase if opinion leaders on social media, friends and family advise so, official guidelines from the Ministry of Health are published and if a GP recommends vaccination”, were also found as significant predictors of the intention to receive COVID-19 vaccine (OR=1.99, 95% CI 1.38–2.87). Susceptibility perceptions (i.e., beliefs concerning the likelihood of someone getting sick from COVID-19 if not getting vaccinated), perceived barriers (time/money) and health motivation (exercise and healthy diet) were not associated with intention to get receive COVID-19 vaccine.

The second model considered in this study, which included TPB variables as well as demographic and health-related factors (Table 4; model 2), explained 64% of the variance in intention to receive COVID-19 vaccine (adjusted R2 = 0.64). The TPB model added 35% to the explained variance on top of the 29% explained by the demographic and health characteristics (Table 4; model 2). According to the TPB model, subjective norms and self-efficacy were significant predictors of intention to get vaccinated against COVID-19. Two subjective norms, namely, “Most of my friends will support the COVID-19 vaccine” and “If I tell my friends and relatives that I intend to get vaccinated against COVID-19 when a vaccine is available, they will respond positively” were significant predictors of intention to receive COVID-19 vaccine (OR=3.04, 95% CI 2.15–4.30). Self-efficacy was also a significant predictor of intention to receive COVID-19 vaccine (OR=2.05, 95% CI 1.54–2.72). PBC and attitude were not significant predictors of intention to receive COVID-19 vaccine.

When key variables from both the HBM and TPB models were entered into a hierarchical regression model (Table 4; model 3), all of the existing relationships remained significant. The combination of HBM and TPB covariates, together with demographic and health-related factors, explained 78% of the variance in intention to get vaccinated against COVID-19 (adjusted R2 = 0.78). Stated differently, adding the TPB covariates on top of the covariates considered in the first model, added 4% to the overall explained variance.

## Discussion

The present study examined the general public’s acceptance and demographical, clinical and psychological predictors of their intention to receive a future COVID-19 vaccine. In examining these predictors, many of this study’s findings were consistent with the results of previous efforts. At the same time, additional predictors of the intention to receive a future COVID-19 vaccine that have not been previously reported, were described here as well.

The overall intention to receive COVID-19 vaccine found in the present study was very high (80%). These results are compatible with the findings of Dror et al., who showed a vaccine acceptance rate of 75% in the entire Israeli population [9], and are also similar to those of Reiter et al. [8], who found that 69% of participants in the United States were willing to receive a COVID-19 vaccine, and to those of Wong et al. [24] showing that 48% of participants definitely intend to receive the vaccine, in addition to the 30% who probably intend to receive the vaccine.

Higher rates were reported among participants aged 65 years and above (93%), similar to earlier work reporting an acceptance rate of 91.3% among Chinese adults [29]. It is reasonable to find higher intention of vaccination among respondents in this age group, as they are also included in the high-risk group for COVID-19. Vaccination intention was lower among several demographic groups, including those aged below 65 years, females and non-academics, similar to what was seen in Fisher et al. and Neumann-Böhme et al., both of which also recommended that in future vaccine programs, efforts be made to target persons below the age of 55 years and at females, and in general, where willingness for vaccination is lower [6, 30].

We examined several predictors that may predict an intention to receive COVID-19 vaccine, which had apparently not been previously reported in the literature in the context of COVID-19 vaccine acceptance. The present study found that respondents with chronic conditions at higher risk of COVID-19 or those over-weight, as well as those who reported having been vaccinated against influenza last year were more likely to accept COVID-19 vaccine. While several other predictors were considered, none were found to be significant in terms of intention to receive COVID-19 vaccine. These included demographic considerations, such as personal status, socio-economic level, residence in the periphery, being an immigrant, number of children and health-related contrasts, such as perceived health status, or having been infected with COVID-19 or influenza in the last year.

Regarding the use of risk perception models, this is apparently the first study to use the TPB model to predict intention to receive COVID-19 vaccine. The theoretical framework at the heart of the present study included demographic variables, health-related factors and both the HBM and TPB risk perception models. This unified model was able to explain 78% of the variance in the intention to receive COVID-19 vaccine. According to HBM, perceived benefits, cues to action, and perceived severity were the most significant predictors of intention to receive COVID-19 vaccine. The findings regarding disease severity indicate that those who intend to get vaccinated view themselves as being at high risk of significant suffering or experiencing complications should they be infected with COVID-19, as compare to those who do not intent to get vaccinated. This indicates the need to increase risk perception and severity among the public, especially among those who perceive the disease as being non-dangerous.

Regarding cues to action, significant predictors which increased the intention to COVID-19 vaccine were recommendations from the Ministry of Health and GP or carrying out the vaccination at the place of work. These observations are similar to findings reported by Reiter et al., who suggested provider recommendation as being a key determinant of vaccination behavior in terms of promoting the vaccine [8]. Regarding the benefits, those who intend to receive the vaccine see high perceived benefits in obtaining the COVID-19 vaccine for protecting themselves and others, similar to what Dror et al. reported, implying that vaccination compliance relies on a personal risk–benefit perception [9]. Finally, according to the TPB model, subjective norms and self-efficacy were significant predictors of intention to get vaccinated against COVID-19. Subjective norms that especially drove respondents were when friends and relatives positively responsed to the vaccination.

It is important to set up intervention plans to deal with respondents with low intentions of receiving the vaccine so as to ensure high actual vaccination uptake, especially among high-risk groups. Hence, public health intervention programs should focus on increasing the perception of vaccination benefits and perceived severity to infection. In addition, investments in the Ministry of Health’s information campaign and resources for providing vaccinations at the workplace should be made.

The COVID-19 epidemic has had an effect not only on the immunization against this disease but also on readiness to receive other vaccines, such as that against influenza. Indeed, a major concern for the coming winter is the combination of COVID-19 and influenza. Previous studies have demonstrated how an influenza pandemic can increase seasonal influenza vaccination acceptance [15], however, it is not clear whether the COVID-19 pandemic has affected influenza vaccine acceptance among the general public. Only few recent studies showed a change in terms of intention to accept seasonal influenza vaccination during the 2019 coronavirus disease pandemic among nurses in Hong Kong, China [29]. Moreover, a study conducted in 17 pediatric emergency departments in 6 countries demonstrated an increase of 15.8% in the number of caregivers who stated they plan to vaccinate their children against influenza, relative to the previous year [31]. The findings of the present study show that the COVID-19 pandemic may have influenced intentions to receive the seasonal influenza vaccinated in the general public. Half of the respondents reported that they had decided not to receive influenza vaccine in the previous season, yet 40% of them indicated that they would be willing to receive influenza vaccine in the coming winter and 66% reported they intend to receive COVID-19 vaccine. While covid-19 vaccination is not yet available, it is also important to increase influenza vaccination rates in the coming winter and deliver that vaccine before the COVID-19 vaccine arrives.

## Limitations

It is important to recognize study limitations when interpreting the results reported here. One limitation of this study is that a convenience sample of participants was recruited via an online survey. Although the demographic characteristics of study participants were similar to those of the general Israeli population, this limitation should be considered in interpreting the results of the study, as our sample population does not include those minorities who do not have high access to online surveys, such as the ultra-Orthodox and Arabs. Additional limitations come from the fact that in the survey used here, vaccination intention was assessed under the assumption that COVID-19 vaccine will be free or covered by basic health insurance. According to Israeli health policy, influenza vaccine is covered by the basic basket of services. Hence, it is reasonable to assume that the COVID-19 vaccine will be similarly covered as part of a budget for preventive services in public health. Finally, the study used self-report of influenza vaccine acceptance in the previous season and intention to influenza vaccine in the coming winter and COVID-19 vaccine once available. Self-report of actual behavior may be biased, unlike monitoring actual vaccination.

## Conclusions

This study provides up-to-date survey data on intention to receive COVID-19 vaccine and highlights the importance of the public perspective. Providing data on the public perspective and predicting COVID-19 vaccination intention by using the HBM and TPB models are important for health policy makers and healthcare providers and can help better guide future compliance once COVID-19 vaccine becomes available. The results highlight that while many adults are willing to get COVID-19 vaccine, vaccination intention differs according to several demographic characteristics. Important predictors of intention to receive COVID-19 vaccine include high perceived benefits, cues to action, and high perceived severity, according to the HBM, as well as subjective norms and self-efficacy, according to the TPB model. Although COVID-19 vaccination is not yet available, it is also important to evaluate intentions to receive influenza vaccine in the coming winter so as to overcome limitations related to behavioral perceptions.

To summarize, the current study examined whether people will accept COVID-19 vaccine as soon as it becomes available. Further research should examine the lag of time of acceptance once such a vaccine is available.

## Data Availability

Not applicable

## Declarations

### Consent for publication

Not applicable

### Availability of data and material

Not applicable

### Authors’ contributions

LS was responsible for study conception and design, data collection and analysis, and wrote the manuscript.

## Acknowledgements

Not applicable

## Author information

Liora Shmueli is a lecturer in the Department of Management, Bar-Ilan University.

